# Concurrent anatomical, physiological and network changes in cognitively impaired multiple sclerosis patients

**DOI:** 10.1101/2020.11.20.20235309

**Authors:** Danka Jandric, Ilona Lipp, David Paling, David Rog, Gloria Castellazzi, Hamied Haroon, Laura Parkes, Geoff Parker, Valentina Tomassini, Nils Muhlert

## Abstract

Cognitive impairment in multiple sclerosis is associated with functional connectivity abnormalities, but the pathological substrates of these abnormalities are not well understood. It has been proposed that resting-state network nodes that integrate information from disparate regions are susceptible to metabolic stress, which may impact functional connectivity. In multiple sclerosis, pathology could increase metabolic stress within axons, damaging the anatomical connections of network regions, and leading to functional connectivity changes. We tested this hypothesis by assessing whether resting state network regions that show functional connectivity abnormalities in people with cognitive impairment also show anatomical connectivity abnormalities.

Multimodal MRI and neuropsychological assessments were performed in 102 relapsing remitting multiple sclerosis patients and 27 healthy controls. Patients were considered cognitively impaired if they obtained a z-score of ≤1.5 on ≥2 tests of the Brief Repeatable Battery of Neuropsychological Tests (n=55). Functional connectivity was assessed with Independent Component Analysis of resting state fMRI images, and anatomical connectivity with Anatomical Connectivity Mapping of diffusion-weighted MRI. Exploratory analyses of fractional anisotropy and cerebral blood flow changes were conducted to assess local tissue characteristics.

We found significantly decreased functional connectivity in the anterior and posterior default mode networks and significant increases in the right and left frontoparietal networks in cognitively impaired relative to cognitively preserved patients. Networks showing functional abnormalities also showed reduced anatomical connectivity and white matter microstructure integrity as well as reduced local tissue cerebral blood flow.

Our results identify key pathological correlates of functional connectivity abnormalities associated with impaired cognitive function in multiple sclerosis, consistent with metabolic dysfunction in functional network regions.

## Introduction

Cognitive impairment is present in up to 70% of people with multiple sclerosis (Chiaravalloti and DeLuca, 2008). Although the disease mechanisms responsible are not fully elucidated (Benedict *et al.*, 2020), cognitive changes in multiple sclerosis are increasingly being viewed as a network dysfunction (Schoonheim *et al.*, 2015*b*; Benedict *et al.*, 2020). Results from resting state functional MRI (rs-fMRI) studies have shown differences in functional connectivity (FC) between cognitively impaired and non-impaired patients. However, the direction of these results is inconsistent, with some studies reporting increased FC in impaired patients and others reporting decreased FC (Hawellek *et al.*, 2011; Loitfelder *et al.*, 2012; Cruz-Gómez *et al.*, 2014; Louapre *et al.*, 2014; Meijer *et al.*, 2017; Rocca *et al.*, 2018). A shortcoming of rs-fMRI, which limits the ability to interpret findings, is the lack of information about pathological mechanisms underlying FC abnormalities.

In Alzheimer’s disease it has been shown that network regions with high connectivity to other parts of the brain, so called ‘hubs’ or ‘central nodes,’ are susceptible to pathological metabolic change (Buckner *et al.*, 2009), and it has been proposed that metabolic stress of network nodes may be responsible for network changes across neurodegenerative diseases (Zhou *et al.*, 2012). This ‘nodal stress hypothesis’ predicts that the high metabolic demands of network nodes makes them susceptible to metabolic stress that accelerates neurodegeneration.

In multiple sclerosis, histological studies have shown changes related to metabolic function in demyelinated axons (Foster *et al.*, 1980; Craner *et al.*, 2004). Metabolic stress within axons could affect the anatomical connections between network regions, either through axonal dysfunction due to a reduced energy state, or through axonal degeneration. Using diffusion MRI (dMRI) we can probe the structural integrity of white matter connecting functional network regions *in vivo*.

Studies which have combined the rs-fMRI and dMRI methods have found a relationship between anatomical and functional abnormalities (e.g. Hawellek *et al.*, 2011; Louapre *et al.*, 2014; Patel *et al.*, 2018; Tewarie *et al.*, 2018). Recently, computational studies support an impact of anatomical connectivity damage on FC. These simulations demonstrated that as white matter pathology initially increased FC increased, but as it continued, FC started to decrease. This finding was interpreted as increased local FC following damage to long-range white matter fibres, followed by a global network collapse with subsequent reductions in FC as the network fails (Patel *et al.*, 2018; Tewarie *et al.*, 2018). Such findings are consistent with the prediction that axonal metabolic stress affects functional networks.

The metabolic stress hypothesis can be further tested by measuring cerebral blood flow (CBF) in functional network nodes. Neural activity requires energy in the form of oxygen and glucose, which are delivered by the blood flow to the brain, and so CBF reflects metabolic demand (Duffin *et al.*, 2018). Hypoperfusion is a well-established finding in multiple sclerosis (Lapointe *et al.*, 2018). and several studies have found links between poor cognitive test performance and reduced CBF or cerebral blood volume (Inglese *et al.*, 2008; D’haeseleer *et al.*, 2013; Francis *et al.*, 2013). Decreased perfusion in multiple sclerosis may be a response to decreased energy demand in affected tissue (Paling *et al.*, 2011; Lapointe *et al.*, 2018) and provides information about the metabolic state of functional network nodes.

In this study we tested the nodal stress hypothesis in a cohort of relapsing-remitting multiple sclerosis patients (RRMS) to determine whether functional network regions that show altered FC also show altered anatomical connectivity and CBF. First, we quantified FC using independent component analysis and dual regression on rs-fMRI data. Second, we applied the Anatomical Connectivity Mapping (ACM) method to characterise anatomical connectivity changes of functional network regions and also assessed fractional anisotropy (FA) to gain information about local white matter tissue characteristics near the network regions investigated. Finally, we assessed cerebral blood flow (CBF), using arterial spin labelling (ASL), within functional network regions.

We predicted that any FC abnormalities in CI patients would co-occur with anatomical connectivity and cerebral blood flow abnormalities.

## Materials and methods

### Participants

One hundred two patients with a diagnosis of RRMS were recruited through the Helen Durham Centre for Neuroinflammation at the University Hospital of Wales. Twenty-seven healthy controls (HC) were recruited from the community. All participants were aged between 18 and 60 years, were right-handed and had no contraindications for MR scanning. Patients had no comorbid neurological or psychiatric disease, were relapse free and had no change to treatment for 3 months prior to undergoing the MRI scan. All participants underwent MRI scanning and assessment of clinical and cognitive function.

The study was approved by the NHS South□West Ethics and the Cardiff and Vale University Health Board R&D committees, and all participants provided written informed consent to participate in the study.

### Clinical and neuropsychological assessment

Clinical functioning was assessed with the Multiple Sclerosis Functional Composite (MSFC), a standardised measure of upper and lower limb and cognitive function (Cutter *et al.*, 1999).

All participants underwent a neuropsychological assessment with the Brief Repeatable Battery of Neuropsychological Tests (BRB-N), a validated testing battery with demonstrated sensitivity to detect cognitive impairment in multiple sclerosis (Amato *et al.*, 2006). The battery includes tests which probe verbal memory (Selective Reminding Test), visuospatial memory (Spatial Recall Test), information processing speed, working memory, attention and executive function (Paced Auditory Serial Addition Test (PASAT) and Symbol Digit Modalities Test), and verbal fluency (World List Generation Test) (Rao, 1990). Patients’ scores on each test were converted to *Z* scores based on means and standard deviations from the 27 HCs. Patients who scored ≥1.5 standard deviations below the control mean, i.e. *Z* ≤ - 1.5, on two or more tests were considered cognitively impaired (CI), a definition of cognitive impairment that is considered to be of medium stringency (Sepulcre *et al.*, 2006). All other patients were considered cognitively preserved (CP). Scores for each of the four cognitive domains of verbal memory, visual memory, attention, information processing and executive function, and verbal fluency, were calculated by averaging the scores for each test in that domain, as described by Sepulcre et al (Sepulcre *et al.*, 2006).

### MRI data acquisition

All participants underwent MRI examination on a 3T MR scanner (General Electric HDx MRI System, GE Medical Devices, Milwaukee, WI) using an eight channel receive-only head RF coil. A high-resolution 3D T1-weighted (3DT1) sequence was acquired for identification of T1-hypointense multiple sclerosis lesions, segmentation, registration and volumetric measurements [resolution 1×1×1 mm, TE = 3.0 ms, TR = 7.8 ms, matrix = 256×256×172, FOV = 256 x 256 mm, flip angle = 20□]. A T2/proton-density (PD)-weighted sequence (voxel size = 0.94×0.94×4.5 mm, TE = 9.0/80.6 ms, TR = 3000 ms, FOV = 240 x 240 mm, 36 slices) and a fluid-attenuated inversion recovery (FLAIR) sequence (voxel size = 0.86×0.86×4.5 mm, TE = 122.3 ms, TR = 9502 ms, FOV = 220 x 220 mm, 36 slices) were also acquired, for identification and segmentation of T2-hyperintense multiple sclerosis lesions. In addition, resting state fMRI was acquired using a T2* weighted gradient-echo echo-planar (GE-EPI) imaging sequence (voxel resolution = 3.4×3.4×3 mm, TE = 35 ms, TR = 3000 ms, FOV = 220 x 220 mm, 100 volumes, 46 axial slices each in an interleaved order). During the fMRI acquisition all participants were instructed to relax with their eyes closed. For the dMRI acquisition, a twice refocused diffusion-weighted spin echo echo-planar (SE-EPI) sequence was acquired with 6 volumes with no diffusion weighting and 40 volumes with diffusion gradients applied in uniformly distributed directions (Camino 40), b = 1200 s/mm^2^, voxel size=1.8×1.8×2.4 mm, TE = 94.5 ms, TR = 16000 ms, FOV = 230 x 230 mm, 57 slices. Finally, CBF was quantified as an indicator of vascular health using multi-inversion time pulsed arterial spin labelling (ASL). A PICORE QUIPSS II sequence with a dual-echo gradient-echo readout and spiral k-space acquisition was employed (voxel size=3×3×8 mm, 22 slices) (Warnert *et al.*, 2015). Sixteen tag-control pairs each for short inversion times, TI (400, 500, 600, 700 ms) and 8 tag-control pairs for long TI (1100, 1400, 1700 and 2000 ms) were acquired with QUIPSS II cut-off at 700 ms. A calibration (M_0_) image was acquired to obtain the equilibrium magnetization of cerebrospinal fluid, needed for the quantification of CBF. A minimal contrast image was acquired with TE=11ms, TR=2000 ms to correct for coil inhomogeneities.

### 3DT1 image analysis

All 3DT1 structural images from patients were lesion filled, to allow better segmentation of brain tissue (Gelineau-Morel *et al.*, 2012). Lesions were identified, binary lesion masks were created and the 3DT1 image was lesion-filled as described by Lipp *et al.*, 2019. The lesion-filled 3DT1 images were segmented into grey matter (GM), white matter (WM) and cerebrospinal fluid (CSF) using FSL’s Automated Segmentation Tool (FAST) (Zhang *et al.*, 2001). The grey and white matter images were binarised and added together to create a binary mask of intracranial brain tissue that excludes CSF, which was used for dMRI analysis. FSL’s SIENAX tool (Smith *et al.*, 2002) was applied to lesion-filled 3DT1 images to quantify brain volumes, including whole brain volume, grey matter volume and white matter volume. All volumetric measurements were normalised for the subject’s intracranial volume. Lesion volume was calculated from the binary lesion masks.

### rs-fMRI analysis

The BOLD fMRI time-series was corrected for physiological noise through a two-step process performed in MATLAB (The MathWorks, n.d.), using a previously established pipeline (Lipp *et al.*, 2014). First, correction of cardiac and respiratory artefacts was applied with RETROICOR tool (Glover *et al.*, 2000), using two cardiac, two respiratory and one interaction components. Second, variance related to CO2 level, O2 level, heart rate and respiration volume per time was regressed out in a general linear model (Birn *et al.*, 2006). Resting state fMRI images were preprocessed with FSL’s MELODIC pipeline (Beckmann *et al.*, 2009), which included motion correction, spatial smoothing with a 3 mm full width at half-maximum Gaussian kernel, high-pass temporal filtering equivalent to 0.01 Hz, non-linear registration to Montreal Neurological Institute (MNI) standard space, and resampling to a resolution of 4 mm isotropic. Head motion parameter estimates of absolute and relative displacement values were not different between any groups (HC-RRMS p=0.58 (absolute), p=0.27 (relative); CP-CI p=0.11 (absolute), p=0.52 (relative)).

Independent Component Analysis (ICA), part of the MELODIC pipeline, decomposed the concatenated dataset into 82 components. Four resting state networks (RSNs) which have been extensively studied and found to be important for cognitive function in multiple sclerosis were selected for further analyses: the default mode network (DMN) (Rocca *et al.*, 2010*b*, 2012; Hawellek *et al.*, 2011; Cruz-Gómez *et al.*, 2014; Meijer *et al.*, 2017), left and right frontoparietal networks (LFPN, RFPN) (Van Den Heuvel *et al.*, 2009; Cruz-Gómez *et al.*, 2014; Louapre *et al.*, 2014; Meijer *et al.*, 2017) and the salience network (SN) (Van Den Heuvel *et al.*, 2009; Rocca *et al.*, 2012; Cruz-Gómez *et al.*, 2014). The anterior and posterior parts of the DMN (DMNa and DMNp, respectively) (Xu *et al.*, 2016), were identified in two additional components and selected for inclusion. The primary visual network was used as a non-cognitive control network, to assess the extent to which FC differences between CI and CP groups are specific to RSNs important for cognition.

Dual regression (Beckmann *et al.*, 2009) was used to generate subject-specific versions of the group-average components. These were fed into FSL’s *randomise* (Winkler *et al.*, 2014) permutation-testing tool, with age, sex and education level as covariates, to assess group differences in FC in the RSNs of interest. We compared both the whole RRMS group with HC, and the CI and CP patient groups to each other. All results were threshold-free cluster enhancement (TFCE) corrected at *p*≤0.05, two-sided. After thresholding, we calculated the percentage of network voxels showing abnormal FC between groups, and retained only those RSNs showing the largest proportion of abnormal network voxels for further analyses. This was done for two reasons. First, we wanted to find pathological substrates of FC abnormalities and chose to investigate the RSNs showing the greatest abnormalities. Second, we wanted to reduce the effect of noise. Rs-fMRI is a noisy method, and while we undertook thorough pre-processing of the data, we can’t be certain that some artifacts aren’t present in our results. Small clusters are more likely to be due to noise (Griffanti *et al.*, 2017) and we therefore focused our further analyses only on those RSNs showing the biggest FC abnormalities in CI patients, in terms of proportion of abnormal network voxels.

### Diffusion MRI processing

Preprocessing of dMRI data was carried out in ExploreDTI (v 4.8.3; (Leemans *et al.*, 2009)) and included corrections for head motion, distortions induced by the eddy currents of the diffusion□weighted gradients and EPI□induced geometrical distortions by registering each diffusion image to its respective (skull stripped and downsampled to 1.5 mm) 3DT1 image (Irfanoglu *et al.*, 2012) using Elastix (Klein *et al.*, 2010), with appropriate reorientation of the diffusion□encoding vectors (Leemans and Jones, 2009). The corrected data were further processed with FSL’s FDT tool to fit diffusion tensors, generate fractional anisotropy (FA) maps and fit the probabilistic diffusion model using the Bedpostx tool (Behrens *et al.*, 2003, 2007).

### Anatomical Connectivity Mapping

ACM uses probabilistic tractography to initiate streamlines from every voxel of the parenchyma, and determines how many pass through each brain voxel, thus assessing the degree of anatomical interconnection of every voxel in the brain (Embleton *et al.*, 2007; Cercignani *et al.*, 2012). ACM maps were generated using FSL’s Probtrackx2 tool (Behrens *et al.*, 2003, 2007). The binary parenchymal mask was used to seed the tractography with 50 initiated streamlines per voxel. The resulting ACM maps show anatomical connectivity across the whole brain, where the magnitude of the ACM value in each voxel represents the number of probabilistic streamlines passing through that voxel (Bozzali *et al.*, 2011). To normalise each ACM for intracranial volume, each individual’s ACM image was divided by the number of voxels in the brain parenchymal mask. To normalise to MNI space, the downsampled 3DT1 image of each participant was nonlinearly registered to MNI space, and the warps were applied to the ACM images. All dMRI parameter maps were analysed in standard (MNI) space.

### ASL image processing

The two sets of ASL tag-control images were first motion corrected to the M0 image using FSL’s McFLIRT tool (Jenkinson *et al.*, 2002), then control-tag subtracted, averaged across pairs, and combined into a single multi-TI series that was fed to *oxford_asl* (BASIL) (Chappell *et al.*, 2009) for CBF quantification. CBF was estimated with partial volume correction (Chappell *et al.*, 2011), coil sensitivity correction (bias field calculated using the SPM12 segmentation (Debernard *et al.*, 2014) on the minimum contrast image) and calibration with the M0 signal from subject-specific ventricle masks. CBF maps were then registered to the T1 structural scan following 6 DOF affine registration of the M0 scan. T1- weighted images were non-linearly normalised to the Montreal Neurological Institute (MNI) 152 template space, using ANTs SyN (Avants *et al.*, 2008) and the obtained warp was applied to the CBF maps. Full CBF maps could not be obtained for all participants due to technical problems with the MR acquisition or due to failed quality checks of the data. CBF analyses were therefore conducted on data from 50 CI and 44 CP patients.

### Statistical analyses

Statistical analyses of the demographic, clinical, global MRI and median ACM, FA and CBF values were performed in SPSS version 23.0 (IMB Corp., 2015). The distributions of all variables were checked through visual inspection of histograms and Q-Q plots, as well as the application of Kolmogorov-Smirnov tests. Variables showing a skew were analysed with non-parametric tests. We then analysed the data in the following steps:

#### 1. Local changes in anatomical connectivity and CBF in regions showing FC changes

We compared ACM, FA and CBF values within those RSN voxels clusters that showed significant FC differences between CI and CP groups. For each RSN a binary mask was created from the voxel clusters that differed in FC between CI and CP patients (referred to as FC-diff masks). The masks (Fig. 1C) were used to extract local median ACM, FA and CBF values from these regions, which were then compared between CI and CP groups.

**Figure 1.**
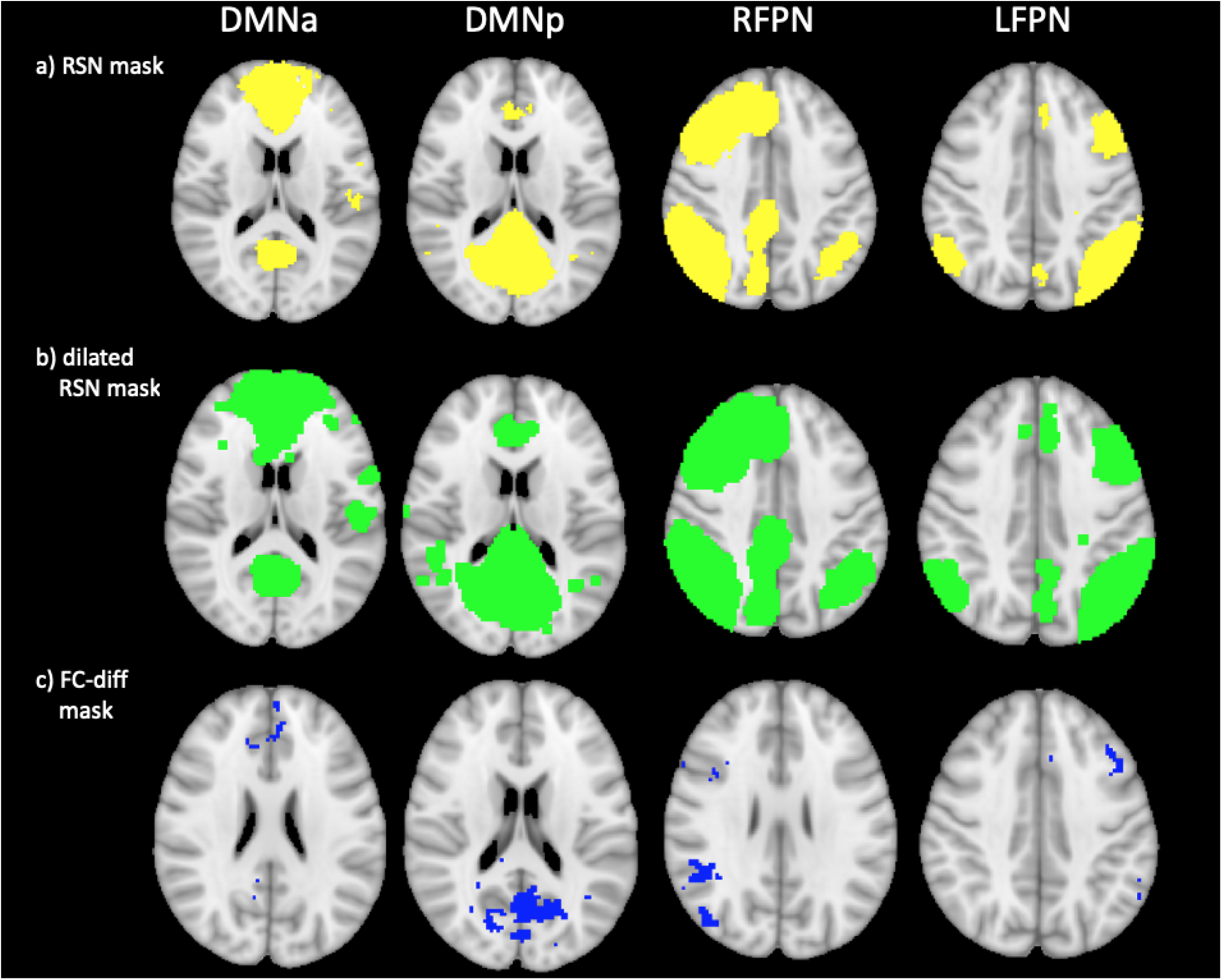
Masks used for dMRI and CBF analyses. Figure 1a shows the binary masks of the RSNs which showed FC abnormalities between CI and CI patients (in yellow). Figure 1b shows the same binary RSN masks dilated by a kernel-width to reach into surrounding white matter for the dMRI analyses (in green). Figure 1c shows the voxel clusters where FC was abnormal in CI compared to CP patients (in blue). All masks are overlaid on a T1-weighted MNI template image.

#### 2. Diffuse changes in anatomical connectivity and CBF within RSNs

Second, we determined whether there were more diffuse changes in anatomical connectivity and CBF throughout each RSN. For this, each individual RSN spatial map was binarised to create a mask (Fig. 1A). For dMRI analyses these binarised masks were dilated by one voxel to include the white matter surrounding RSN regions (Fig. 1B). The masks were used in two ways. First, we ran voxelwise analyses of ACM, FA and CBF maps to look for clusters of abnormalities within the RSN regions. For FA, this was done both with skeletonised FA maps in a tract-based spatial statistics (TBSS) analysis (Smith *et al.*, 2006), and with non- skeletonised FA maps in an exploratory analysis. Next, in additional exploratory analyses we extracted median ACM, FA and CBF values from the RSN regions to determine differences within RSNs between CI and CP patients. All exploratory analyses are described in the Supplementary material.

#### 3. Diffuse changes in anatomical connectivity and CBF throughout the brain

Last, we checked whether CI and CP groups showed differences in ACM, FA and CBF throughout the brain, by running voxelwise analysis on the ACM, FA and CBF maps of the whole brain. This was an exploratory analysis to understand the extent of ACM, FA and CBF changes in CI patients relative to CP patients, and is described in the Supplementary material.

#### Comparisons, thresholding and multiple comparison correction

Comparisons of FC changes were conducted for both the whole RRMS group with HC, and the CI and CP patient groups to each other. While the comparison of the two patient groups was based on our hypotheses, a comparison of the whole group to controls would let us determine the extent to which any detected FC abnormalities were specific to patients with cognitive impairment. However, subsequent analyses, of the anatomical connectivity and cerebral blood flow, were conducted only for the two patient groups to limit the number of statistical comparisons and in line with our hypotheses which focus on the differences between cognitively impaired and non-impaired patients.

Comparisons of median ACM, FA values and CBF values were performed using a two-sample t-test or Mann Whitney U test, as appropriate. A Bonferroni correction for multiple comparisons, of a factor of four for the four RSNs of interest, was applied to the results. The corrected threshold was *p*≤0.0125.

Age, sex and education level were included in general linear models for all voxelwise analyses as covariates, and all results were TFCE-corrected at *p*≤0.05, two-sided. The Harvard-Oxford cortical structural, Harvard-Oxford subcortical structural and JHU white-matter tractography atlases in FSL were used to report anatomical locations.

## Data availability

Data are available from the corresponding authors upon reasonable request.

## Results

### Demographic, clinical, neuropsychological characteristics and conventional MRI data

The demographic, clinical and neuropsychological characteristics of healthy controls, CI and CP patients, as well as the whole patient group, are presented in Table 1.

**Table 1.** Demographic, clinical and neuropsychological characteristics

|  | **HC**<br>**(n=27)** | **RRMS**<br>**(n=102)** | *Inferential test*<br>*results*<br>*HC-RRMS*<br>*comparisons* | **RRMS CP**<br>**(n=47)** | **RRMS CI**<br>**(n=55)** | *Inferential test results*<br>*CI-CP comparisons*<br>*HC-CI-CP comparison*<br>*for BRB-N Z-scores* |
| --- | --- | --- | --- | --- | --- | --- |
| **Demographic characteristics** |  |  |  |  |  |  |
| **Age, yr (median,**<br>**range)** | 37.00 (23-59) | 45.00 (18-60) | $U = 958.00, p = .015$ | 42.00 (18-60) | 47.00 (20-60) | $U = 1069.50, p = .134$ |
| **Male/female, n** | 12/15 | 33/69 | $\chi^2(1) = 1.37, p = .241$ | 11/36 | 22/33 | $\chi^2(1) = 3.19, p = .074$ |
| **Education years**<br>**(median, range)** | 19.00 (12-30) | 15.00 (10-30) | $U = 613.50, p < 0.001$ | 15.00 (10-27) | 14.00 (10-30) | $U = 1084.50, p = .161$ |
| **Mean disease**<br>**duration, yr**<br>**(median, range)** | N/A | 12.24 (1-39) | N/A | 11.50 (2-37) | 12 (1-39) | $U = 1232.50, p = .803$ |
| **MSFC** |  |  |  |  |  |  |
| **25 Foot Walk**<br>**Test (median,**<br>**range)** | 4.35 (3.2-5.4) | 5.25 (3.6-26.8) | $U = 572.50, p < 0.001$ | 5.15 (3.7-13.0) | 5.43 (3.6-26.8) | $U = 1169.50, p = .498$ |
| **9-Hole Peg Test**<br>**(median, range)** | 18.65 (15.35-23.00) | 21.75 (16.35-59.50) | $U = 537.50, p < 0.001$ | 21.45 (17.15-44.85) | 21.95 (16.35-59.5) | $U = 956.00, p = .024$ |
| **PASAT3**<br>**(median, range)** | 51.00 (35-59) | 43.50 (0-60) | $U = 715.00, p < 0.001$ | 50.00 (30-60) | 34.00 (0-58) | $t(83.10) = 6.50, p < 0.001$ |
| **BRB-N Z-scores** |  |  |  |  |  |  |
| **Verbal memory**<br>**(mean, SD)** | 0.00 (0.92) | * | N/A | 0.07 (0.071) | -1.53 (1.09) | $F(2, 125) = 44.82, p < .001$<br>Post hoc:<br>HC-CI $p < .001$<br>HC-CP $p = .931$<br>CP-CI $p < .001$ |
| **Visual Memory**<br>**(mean, SD)** | 0.00 (0.92) | * | N/A | -0.13 (0.93) | -1.20 (1.01) | $F(2, 126) = 22.36, p < .001$<br>Post hoc: |
| | | | | | | HC-CI $p < .001$<br>HC-CP $p = .883$<br>CP-CI $p < .001$ |
| Information processing, attention, executive function (mean, SD) | 0.00 (0.75) | * | N/A | -0.37 (0.73) | -1.90 (1.26) | $F(2, 126) = 44.58, p < .001$<br>Post hoc:<br>HC-CI $p < .001$<br>HC-CP $p = .298$<br>CP-CI $p < .001$ |
| Verbal fluency (mean, SD) | 0.00 (1.00) | * | N/A | 0.08 (0.72) | -0.51 (0.93) | $F(2, 125) = 6.33, p = .002$<br>Post hoc:<br>HC-CI $p = .04$<br>HC-CP $p = .932$<br>CP-CI $p = .003$ |
| **MRI volume measures** |  |  |  |  |  |  |
| Normalised brain volume, L (median, range) | 1.55 (1.42-1.70) | 1.55 (1.42-1.70) | $t(41.94) = 3.33, p = .002$ | 1.50 (1.37- 1.66) | 1.51 (1.30- 1.68) | $t(99.83) = 0.36, p = .721$ |
| Normalised grey matter volume, L (median, range) | 0.81 (0.72- 0.89) | 0.77 (0.61- 0.89) | $U = 755.00, p < 0.001$ | 0.77 (0.61- 0.89) | 0.76 (0.62- 0.88) | $t(99.83) = 1.48, p = .142$ |
| Normalised white matter volume, L (median, range) | 0.76 (0.68- 0.81) | 0.74 (0.66- 0.83) | $t(40.43) = 1.56, p = .127$ | 0.74 (0.66- 0.81) | 0.75 (0.66- 0.83) | $t(97.31) = -1.24, p = .218$ |
| Lesion volume, mL (median, range) | N/A | * | | 9.73 (0.64- 63.32) | 9.73 (0.69- 59.64) | $U = 1258.00, p = .817$ |

Independent samples t-tests were used for group comparisons of variables with a normal distribution. Those variables which were not normally distributed were assessed with Mann-Whitney U tests. Categorical variables were tested with the chi-squared test. BRB-N Z-scores were tested with a one-way ANOVA and Tukey's post hoc test. \*RRMS group averages not calculated.
Abbreviations: BRB-N = Brief Repeatable Battery of Neuropsychological Tests; CI = cognitively impaired; CP = cognitively preserved; HC = healthy controls; MSFC = Multiple Sclerosis Functional Composite; PASAT3 = Paced Auditory Serial Addition Test; RRMS = relapsing remitting multiple sclerosis; SD = standard deviation

RRMS patients and controls showed no significant differences in sex, but the RRMS group was significantly older and less educated than controls and performed worse on all MSFC tests.

Fifty-five patients met the definition for cognitive impairment, and 47 were considered cognitively preserved. Compared to CP patients, the CI group did not differ significantly in age, education, disease duration, proportion of males and females, or lower limb function, as measured by the 25 Foot Walk Test of the MSFC. However, their performance on the 9-Hole Peg Test was slower, demonstrating worse upper limb function, and as expected, they scored significantly worse on the PASAT than the CP group. Similarly, CI patients showed impaired cognitive function compared to CP patients and HC on all four cognitive domains assessed by the BRB-N. The greatest impairments were observed on the information processing, attention and executive function (*z* score = -1.90) and verbal memory domains (*z* score = - 1.53). Cognitively preserved patients did not perform significantly worse than controls on any domain. Full results are presented in Table 1 below.

The patient group had significantly lower normalised brain volume (NBV) and normalised GM volume (NGMV) than healthy controls, but showed no significant difference in normalised WM volume (NWMV). CI and CP groups showed no significant differences in any of these measures or lesion volumes (Table 1 and Supplementary Fig. 1).

### Functional connectivity

RRMS patients had areas of decreased FC in the salience network and bilateral FPN compared to HC. RRMS also showed increased FC compared to HC, in areas of the DMNa, DMNp, SN, bilateral FPN, and the primary visual network (Fig. 2).

**Figure 2.**
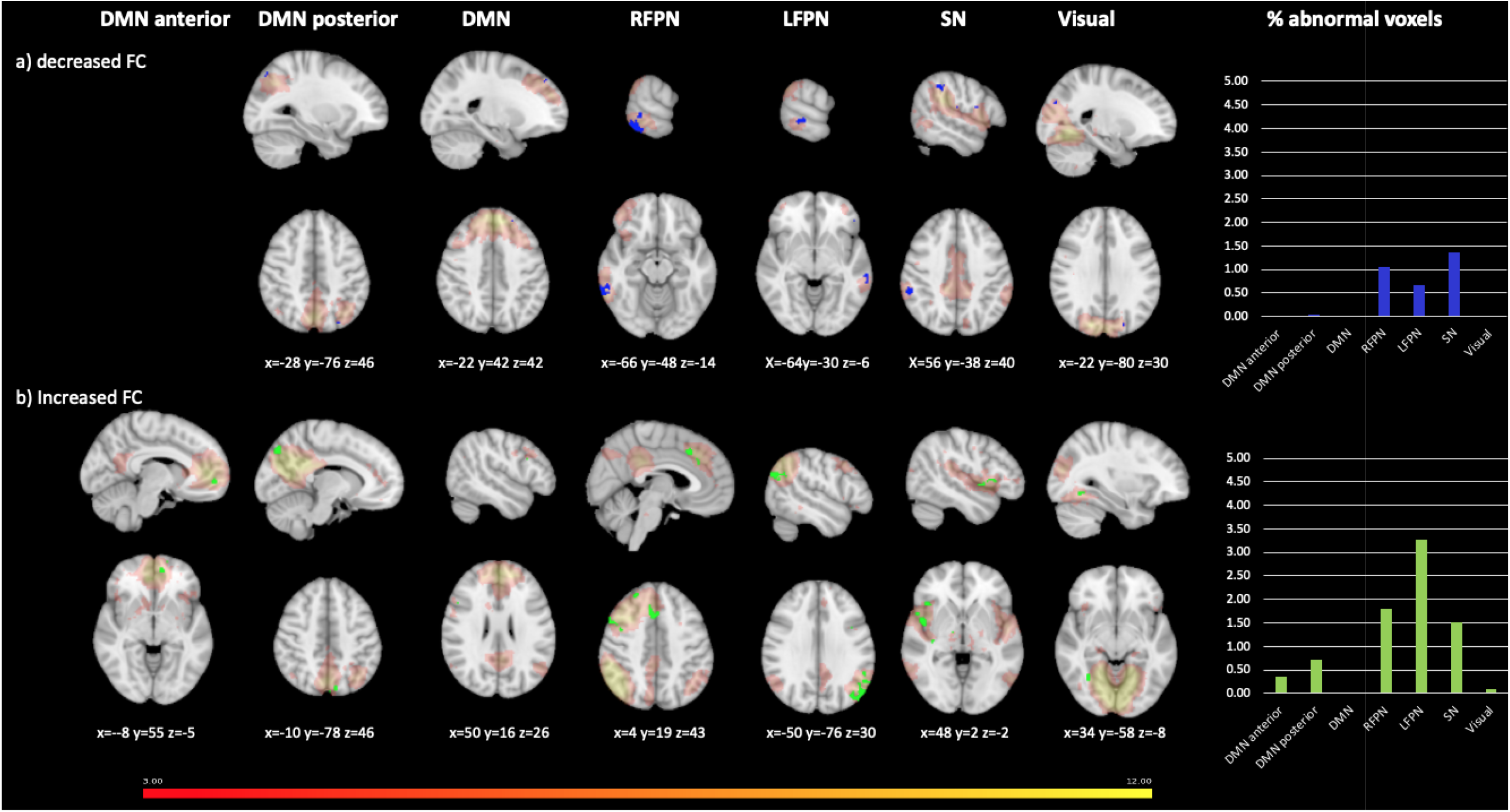
Functional connectivity abnormalities in the RRMS group compared to healthy controls. Figure shows voxels showing FC abnormalities in RRMS compared to healthy controls, overlaid onto the group average spatial map of each RSN analysed in red-yellow. First seven columns show each of the RSNs investigated: DMN anterior, DMN posterior, DMN, RFPN, LFPN, SN and primary visual network. For networks not displayed, no significant group differences were found. The eighth column shows graphs indicating the percentage of voxels showing abnormalities, of the total number of voxels in the network. Rows show areas of: a) decreased FC in the RRMS group vs HC (in blue); b) increased FC in RRMS group vs HC (in green). Results were TFCE-corrected at *p*≤0.05, two-sided. MNI coordinates are given for results displayed. Colour bar shows signal intensity of RSNs.

When comparing the CI and CP groups (Fig. 3), decreases in the CI group relative to the CP group were found in the DMNa, DMN, DMNp, LFPN and primary visual network. Increases in FC in CI compared to CP patients were observed in the DMN, SN, RFPN, LFPN and primary visual network. Of these, the DMNa, DMNp, LFPN and RFPN showed the largest proportion of abnormal voxels between groups, and were therefore retained for subsequent analyses.

**Figure 3.**
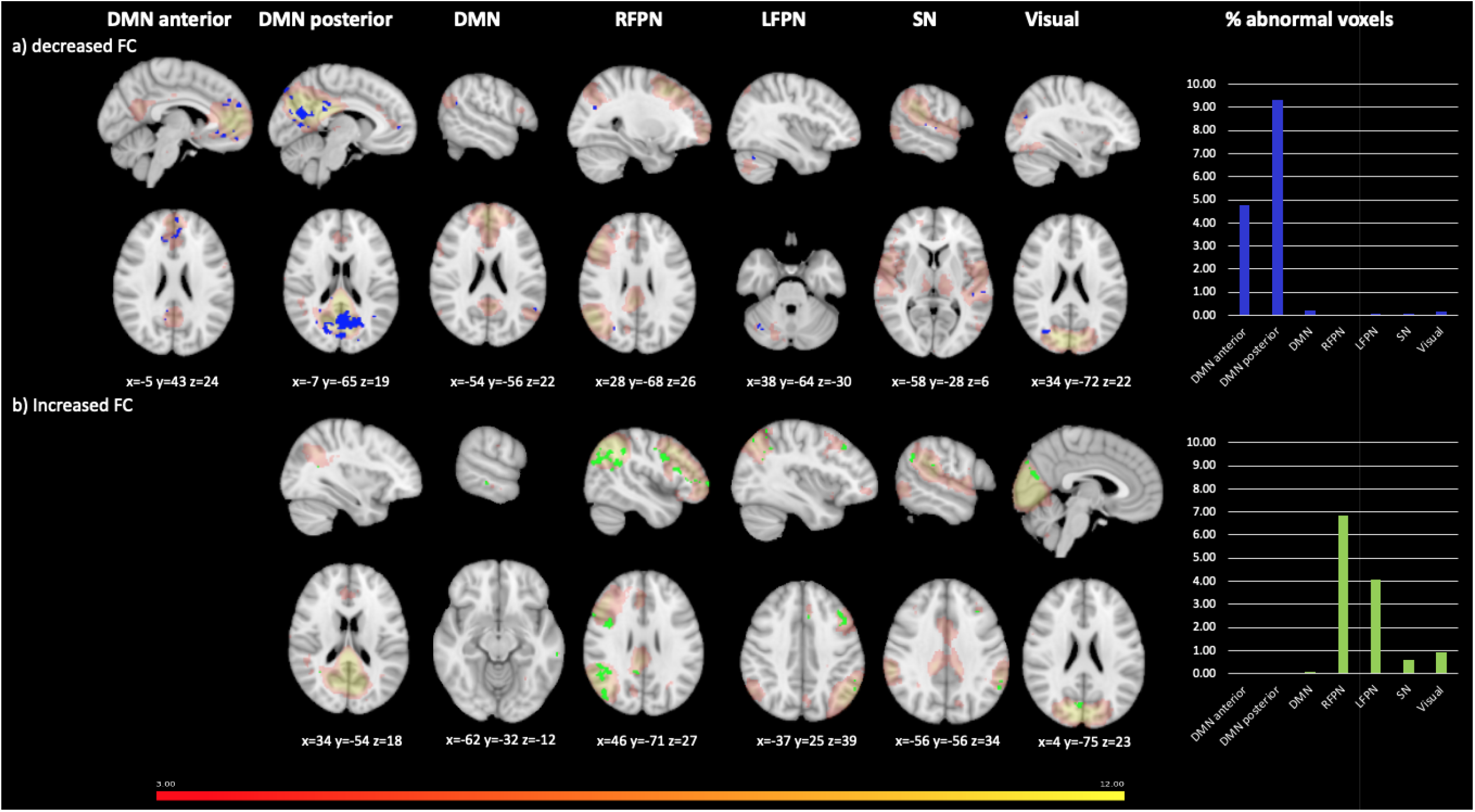
Functional connectivity abnormalities in cognitively impaired compared to cognitively preserved patients. Figure shows voxels showing FC) abnormalities in CI compared to CP, overlaid onto the group average spatial map of each RSN analysed in red-yellow. First seven columns show each of the RSNs investigated: DMN anterior, DMN posterior, DMN, RFPN, LFPN, SN and primary visual network. For networks not displayed, no significant group differences were found. The eight column shows graphs indicating the percentage of voxels showing abnormalities, of the total number of voxels in the network. Rows show areas of: a) decreased FC in the CI group vs CP (in blue); b) increased FC in CI group (in green). Results were TFCE-corrected at *p*≤0.05, two-sided. MNI coordinates are given for results displayed. Colour bar shows signal intensity of RSNs.

### Anatomical connectivity and cerebral blood flow

#### 1. Local changes in ACM, FA and CBF in regions showing FC changes

A comparison of median ACM values, normalised for head size, median FA values and median CBF values, between the CI and CP groups, showed no significant group differences following application of a Bonferroni correction for multiple comparisons (corrected *p* threshold = 0.0125), in the regions of the RSNs that showed FC changes in CI patients compared to CP patients (Supplementary Fig. 2).

#### 2. Diffuse changes in connectivity within RSNs

The voxelwise analysis of ACM maps showed regions of reduced ACM values in all four networks tested (Fig. 4) in CI compared to CP patients. In the DMNa reductions were in regions that correspond to the forceps minor, left cingulum, left anterior thalamic radiation and right anterior corona radiata. In the DMNp reductions were seen in parts of the splenium of the corpus callosum, left and right cingulum, forceps major and also forceps minor. In the RFPN reductions were mainly in white matter corresponding to parts of the right inferior longitudinal fasciculus (ILF) and the right inferior fronto-occipital fasciculus (IFOF). Finally, in the LFPN reductions were in parts of the left superior longitudinal fasciculus, left ILF and left side of forceps major.

**Figure 4.**
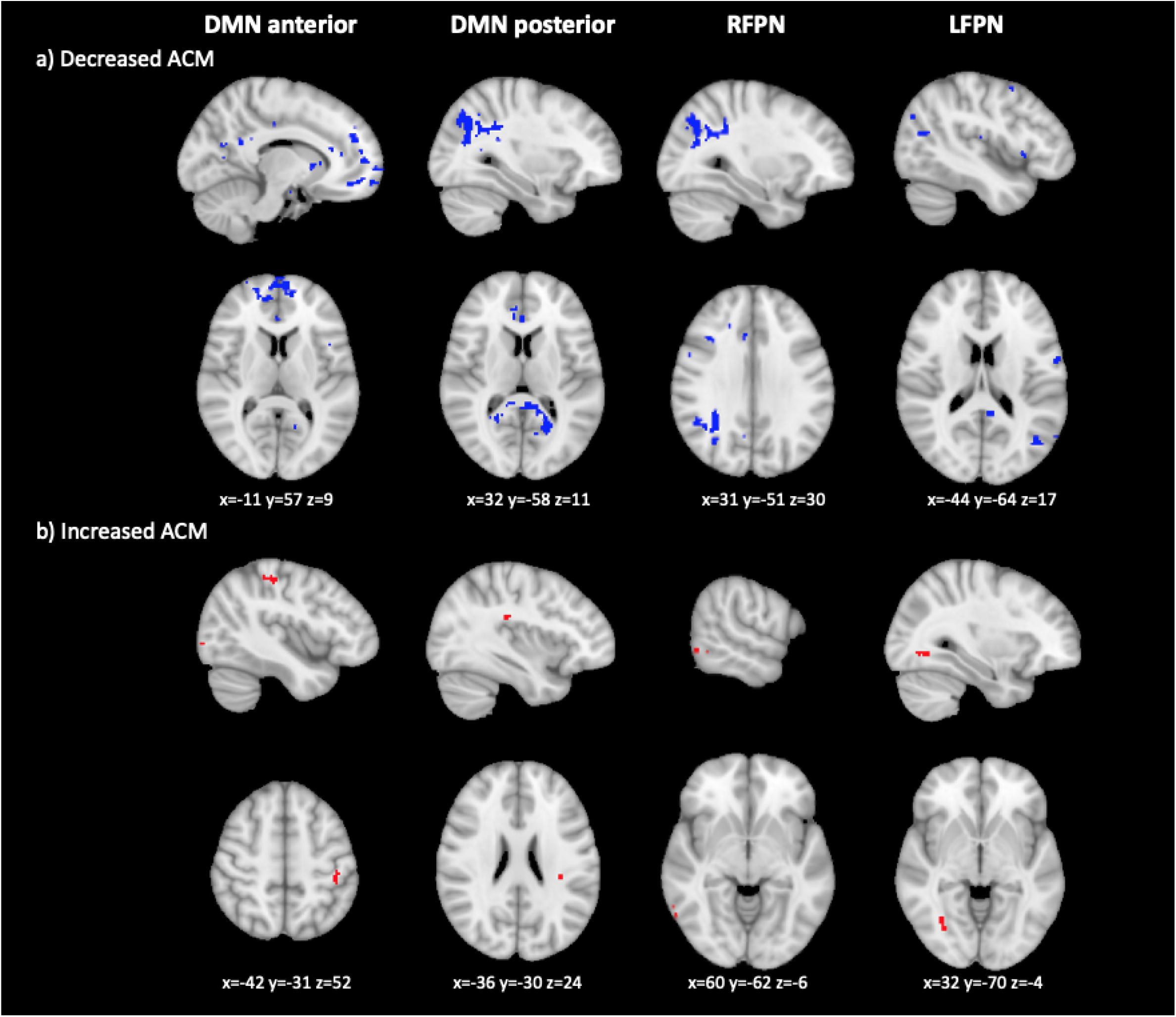
Anatomical connectivity changes in CI compared to CP patients, based on a voxelwise analysis of anatomical connectivity maps. Figure shows voxels showing ACM value abnormalities. Columns show each of the RSNs compared. The first row shows areas of decreased ACM values (in blue), the second row areas of increased ACM values (in red). MNI coordinates are given for the biggest voxel clusters displayed. Results were TFCE-corrected at *p*≤0.05, two-sided.

There were some areas of increased ACM values, including some voxels in the left superior parietal lobe and left occipital lobe in the DMNa, in a part of the left superior longitudinal fasciculus in the DMNp, the right posterior temporal lobe in the RFPN and in regions of the occipital lobe which could be in either the right ILF or right IFOF in the LFPN (Fig. 4).

The TBSS analysis showed reduced FA in CI compared to CP in all four networks investigated. Within the DMNa FA reductions were observed mainly in the genu of the corpus callosum, forceps minor and cingulum bilaterally. In the DMNp FA reductions were in the splenium of the corpus callosum, posterior parts of the cingulum bilaterally and posterior corona radiata bilaterally. In the RFPN there was reduced FA in parts of the right frontal lobe and right parietal lobe, and in the LFPN in the left side of the splenium of the corpus callosum, left side of forceps major and left cingulum.

There were also small areas of FA increases, across the frontal and parietal lobes (Fig. 5).

**Figure 5.**
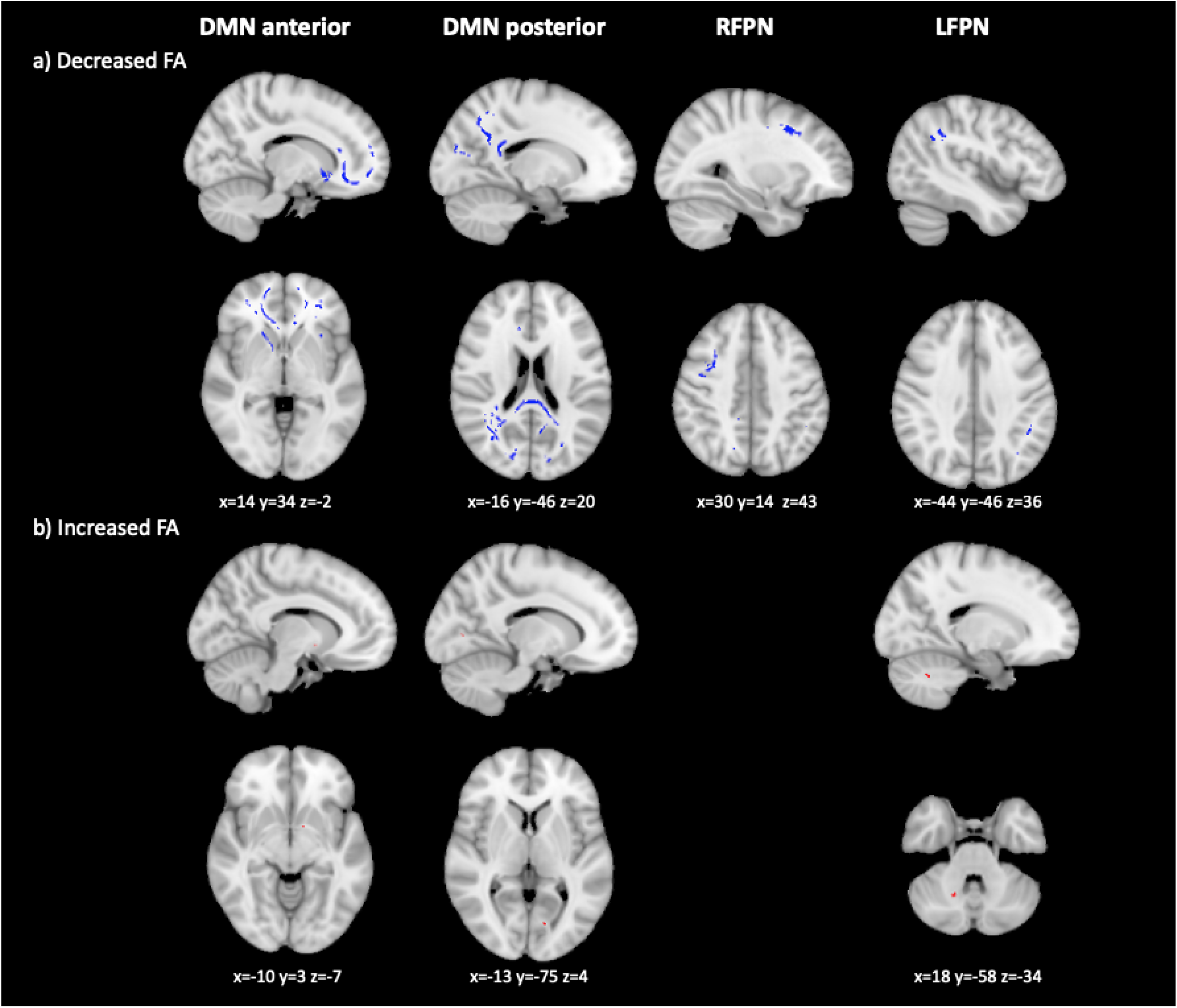
FA changes in CI compared to CP patients, based on a TBSS analysis. Figure shows voxels showing FA abnormalities. Columns show each of the RSNs compared. The first row shows areas of decreased FA (in blue), the second row areas of increased FA (in red). MNI coordinates are given for the biggest voxel clusters displayed. For networks not displayed, no significant results were found. Results were TFCE-corrected at *p*≤0.05, two-sided.

The analysis of CBF maps (Fig. 6) showed widespread regions of reduced CBF in all four network in CI, compared to CP patients. Reductions were seen in the bilateral cingulate gyrus and precuneus in the DMNa; bilateral precuneus, left cuneal cortex, right lateral occipital cortex, left lingual gyrus and left posterior cingulate gyrus in the DMNp; the right occipital cortex, right angular gyrus, right superior supramarginal gyrus and right cingulate gyrus in the RFPN. The same regions but in the left hemisphere showed CBF reductions in the LFPN. We found some individual voxels, likely artefacts, showing increased CBF in CI patients, in the DMNa, DMNp and RFPN.

**Figure 6.**
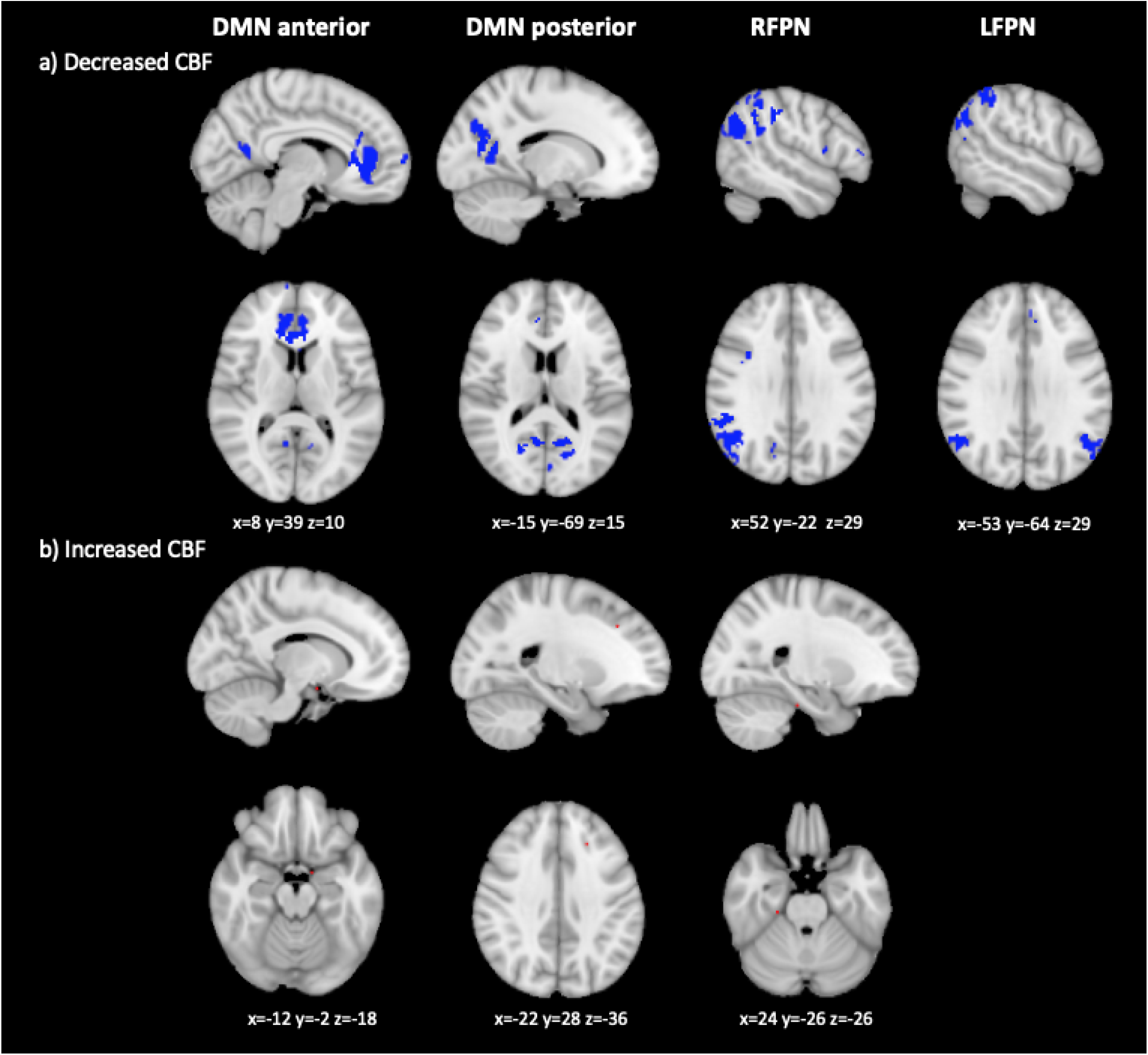
CBF changes in CI compared to CP patients, based on a voxelwise analysis of CBF maps. Figure shows voxels showing CBF abnormalities in red. Columns show each of the RSNs compared. The first row shows areas of decreased CBF (in blue), the second row areas of increased CBF (in red). MNI coordinates are given for the biggest voxel clusters displayed. Results were TFCE-corrected at *p*≤0.05, two-sided.

## Discussion

In this study we tested the ‘nodal stress’ hypothesis (Zhou *et al.*, 2012) as a potential explanation of abnormal resting state FC changes in cognitively relevant functional networks. We looked at FC changes in six RSNs; the DMN and the anterior and posterior DMNs, the left and right FPN, and the SN. In addition, we included the primary visual network as a control region for assessing differences between CI and CP patients. We expected to find FC abnormalities in these networks in RRMS patients with cognitive impairment, and further, that we would see anatomical connectivity abnormalities, assessed through the ACM method, in the networks that showed FC differences in CI compared to CP patients. We further included the FA and CBF measures of white matter integrity and blood perfusion, respectively, to assess the local tissue characteristics of RSN regions in exploratory analyses. We found a pattern of FC decreases and increases, along with ACM, FA and CBF abnormalities, in the DMNa, DMNp, RFPN and LFPN. Thus, our results show that functional networks which show FC abnormalities also show structural abnormalities and altered local tissue characteristics, consistent with metabolic changes resulting in FC abnormalities, as predicted by the nodal stress hypothesis.

### Functional connectivity

Comparing the RRMS group with HC on the FC measure allowed us to determine whether the multiple sclerosis group showed FC abnormalities, as per previous studies (Rocca *et al.*, 2010*b*, 2012, 2018; Roosendaal *et al.*, 2010; Faivre *et al.*, 2012; Loitfelder *et al.*, 2012; Janssen *et al.*, 2013; Koenig *et al.*, 2013; Basile *et al.*, 2014; Tona *et al.*, 2014; Sbardella *et al.*, 2015, 2017; Hulst *et al.*, 2015). We found FC abnormalities in RRMS patients in all RSNs investigated, confirming FC changes as a pathological feature of multiple sclerosis.

A comparison of CI and CP patient groups enabled us to identify how FC relates to cognitive dysfunction in our sample. We found FC abnormalities in the CI group in all networks investigated, with the DMNa, DMNp, RFPN and LFPN showing the highest proportion of affected network voxels and therefore being retained for further analyses. These results are consistent with numerous previous reports of abnormal FC in these networks in patients with cognitive symptoms (Hawellek *et al.*, 2011; Cruz-Gómez *et al.*, 2014; Louapre *et al.*, 2014, Rocca *et al.*, 2016*a*; Meijer *et al.*, 2017; Eijlers *et al.*, 2019; Manca *et al.*, 2019).

In our sample FC was decreased in the DMNa and DMNp and increased in the bilateral FPN in CI relative to CP patients and it is important to consider the potential pathology that could underpin these abnormalities. Past studies have interpreted FC changes in terms of functional re-organisation, that can be either adaptive or maladaptive (Rocca *et al.*, 2010*a*, 2016*b*; Hawellek *et al.*, 2011; Loitfelder *et al.*, 2012; Cruz-Gómez *et al.*, 2014; Tona *et al.*, 2014, Schoonheim *et al.*, 2015*b, a*), but the FC measure does not in itself contain any information about the neural basis of the signal, and so these interpretations are speculative. Strictly speaking, an FC change is simply a change in the level of coherence of the affected voxels with the rest of the voxels in the network (Tahedl *et al.*, 2018). Recent studies have been combining rs-fMRI with other MR modalities to understand FC changes better, and emerging evidence suggests that the direction of FC changes reflects structural damage (Meijer *et al.*, 2018; Patel *et al.*, 2018; Tewarie *et al.*, 2018). Both Patel et al 2018 and Tewarie et al 2018 showed, in their computational modelling studies, that early disconnection mechanisms in the white matter induce increases in functional network connectivity, followed by decreases as structural damage accumulates. It was suggested that a pattern of initial increases in FC followed by decreases could be evidence of damage to long-range white matter tracts connecting RSNs to the rest of the brain, followed by a more global collapse of functional networks. Applying these conclusions to our results would suggest that the DMNa and DMNp, which had reduced FC in CI patients, have sustained more damage than the RFPN and LFPN, which showed increased FC.

### Anatomical Connectivity

Our methods were not designed to test the stage of structural damage, but we did assess the presence of damage in or around RSN regions. For this we used the ACM metric, a network measure of anatomical connectivity that shows how well-connected a specific region is to the rest of the brain. ACM does not require *a priori* assumptions of where in the brain WM damage has occurred, but allows us to ask if anatomical connectivity of a region of interest is affected as a result of WM damage somewhere in the brain.

We found ACM reductions in all four RSNs, highlighting reduced anatomical connectivity of networks showing functional connectivity abnormalities in CI patients. Critically, the specific voxels showing FC abnormalities were not those which showed structural or physiological changes. Instead, other parts of the RSNs were affected. This, combined with widespread anatomical connectivity changes in CI relative to CP patients, suggests more diffuse, as opposed to focal changes within RSNs are associated with cognitive impairment.

### Exploratory analyses of FA and CBF

While the ACM method was chosen for its ability to assess anatomical connectivity, it is a network measure and the information it provides about RSN regions is limited to how well-connected those regions are to other parts of the brain. To better understand local tissue characteristics of RSN regions, we complemented it with the more conventional diffusion metric of FA. FA is a measure of the directionality of diffusion within tissue, which is assumed to be largely determined by the presence of aligned axons in white matter bundles (Beaulieu, 2002). It can therefore give information about local microstructural integrity in a white matter tract and help us understand local tissue characteristics of RSN regions. If there are metabolic changes in the WM in and around RSN regions, the FA metric should be affected. In addition, because it has previously been observed that perfusion in multiple sclerosis is abnormal and may be a response to decreased energy demand in affected tissue (Paling *et al.*, 2011; Lapointe *et al.*, 2018), we assessed CBF. FA and CBF metrics were investigated in exploratory analyses, to gain further information about local WM changes in RSN regions and the energy demands of these regions, respectively.

In CI patients we found regional FA and CBF abnormalities, with reductions in and around RSN regions, if not within the specific voxel clusters showing FC abnormalities. This points to diffuse local tissue abnormalities throughout RSNs. CBF reductions could reflect a response to decreased energy demand in the affected tissue, which supports the key premise we are investigating, of metabolic damage in functional network regions. But it is important to consider that CBF is not a direct measure of lowered neuronal energy demand, and there are suggestions that it could instead be evidence of a primary vascular insult (Lapointe *et al.*, 2018). Future studies with more direct measures of metabolism could help elucidate the metabolic status of RSNs.

### ACM and FA increases

We found small regions of increased ACM and FA in all four RSNs, which are not well supported by the literature. One possibility is that these are statistical artefacts. ACM increases could reflect an unmasking effect, whereby tracking becomes easier in regions where fibres are lost. However, these findings could also reflect plasticity. Bozzali *et al.* (2011) found ACM increases in Alzheimer’s patients and considered that they may be due to plasticity driven by medication. We have not analysed the medication status of our cohort and therefore cannot speculate on this possibility, which will be an important avenue for future research.

The mechanism of FA increases is not well understood, but it has been suggested that increased FA may reflect changes in axonal structures such as reduced branching, decreased axon diameter, reduced packing density, or increases in myelination (Beaulieu, 2002; Hoeft *et al.*, 2007). In MS, FA increases have been considered to be related to inflammatory processes (Calabrese *et al.*, 2011). We can not conclude which mechanism is responsible for the FA increases in our CI group, but acknowledge the finding as a likely important pathological process which should be investigated in future work.

### Methodological considerations

Our approach of combining several MR modalities to investigate the mechanisms of FC changes is relatively novel, and as such we conducted several exploratory analyses to gain an understanding of how best to explore changes in WM metrics and CBF in and around functional network regions. Specially, we investigated ACM, FA and CBF changes using both voxelwise analyses and by looking at average values of each metric in our regions of interest. Furthermore, we used both TBSS and non-skeletonised FA maps for the voxelwise analysis of FA. The reasons for these choices are outlined in the Supplementary material.

From our results we conclude that the voxelwise analysis is more sensitive to group differences than the extraction and comparison of average values in a region, given that the former showed significant group differences and the latter did not. This lack of significant group differences in average values points to heterogeneity in the metrics tested across the regions of interest, which an extraction of average value cannot reflect.

The TBSS analysis and the voxelwise analysis of non-skeletonised FA maps both showed FA reductions in largely the same regions. The latter additionally showed FA changes at the white-grey matter boundaries, which could reflect either partial volume effects due to inaccurate registration of the white matter, or genuine group differences in the grey matter, as has been reported in several studies (reviewed in Inglese and Bester, 2010). However, the finding of group differences at the edge of the brain and at the midline with the voxelwise analysis of non-skeletonised FA images points to registration problems, highlighting that TBSS is a more suitable approach.

### Limitations and future directions

It must be acknowledged that we used several exploratory analyses in this study, the results of which should be interpreted with caution. Further, when interpreting the results it is important to also bear in mind the limits of the methods used to probe metabolic changes. Our results support our prediction that that if network dysfunction is driven by axonal metabolic stress it may be possible to see this on measures of white matter integrity, like FA or ACM, or on measures of cerebral blood flow, they are indirect measures of metabolic changes. Other MR modalities, such as 23Na MRI, can show changes in sodium concentration in tissue, which is a measure of the energy state of axons (Paling *et al.*, 2011). 23Na MRI has been used to investigate cognitive impairment in multiple sclerosis (Paling *et al.*, 2013; Maarouf *et al.*, 2014, 2017) and, if combined with rs-fMRI, has the potential to be informative about the metabolic basis of FC changes.

### Summary and conclusion

In summary, we showed that functional connectivity abnormalities in cognitively impaired RRMS patients co-occur with reduced anatomical connectivity and abnormalities in the cerebral blood flow and local white matter microstructure of network regions. These findings inform us about the pathological correlates of functional connectivity changes which are associated with cognitive impairment, and point to potential metabolic stress to network regions, as per the nodal stress hypothesis. The metabolic state of functional networks affected by multiple sclerosis should be further investigated with more direct methods of metabolic brain function, such as 23Na MRI, to determine the pathological basis of functional connectivity abnormalities.

## Data Availability

Data are available from the corresponding authors upon reasonable request.

## Acknowledgements

The authors would like to thank Dr Daniele Mascali and Dr Antonio Maria Chiarelli at the Gabriele d’Annunzio University of Cheti and Pescara, Italy, for computing the CBF maps used in this study.

## Funding

This work was funded by grants held by VT, IL and DJ. IL and VT were supported by a grant from MS Society UK. DJ was supported by a Medical Research Council Doctoral Training Partnership grant.

## Competing interests

The authors report no competing interests.

## Supplementary material

Supplementary material is available as a separate file.

## Notes

### Competing Interest Statement

The authors have declared no competing interest.

### Author Declarations

NHS South-West Ethics and the Cardiff and Vale University Health Board R&D committees.

### Summary of Updates

CBF data re-analysed following a correction to CBF calculation. Changes affect ASL processing subsection of the methods section, CBF results, figure 6 in the main paper and figures 2,3 and 5 in supplementary materials.

