## Supplementary material for "Concurrent anatomical, physiological and network changes in cognitively impaired multiple sclerosis patients"

### Conventional MRI data analysis - results

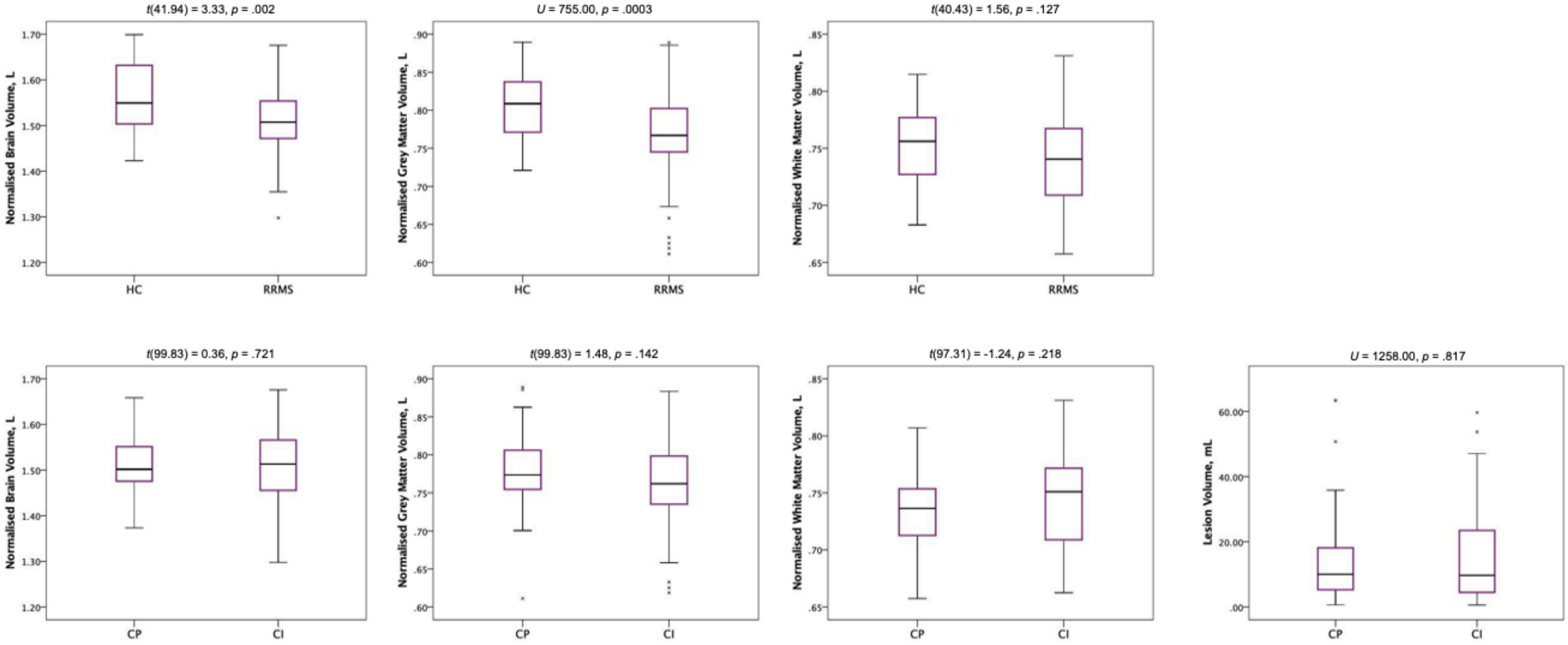

### Figure 1. Group comparisons of structural MRI volume measures

NBV, NWMV and NGMV in the HC and RRMS groups are reported in the top row, and the same measures and measures of lesional volume are shown for CI and CP patients in the bottom row. Boxes show median and interquartile range (IQL), and whiskers show range of values, excluding outliers. Stars indicate outliers.

**Local changes in ACM, FA and CBF in regions showing FC changes – method and results**

**A**

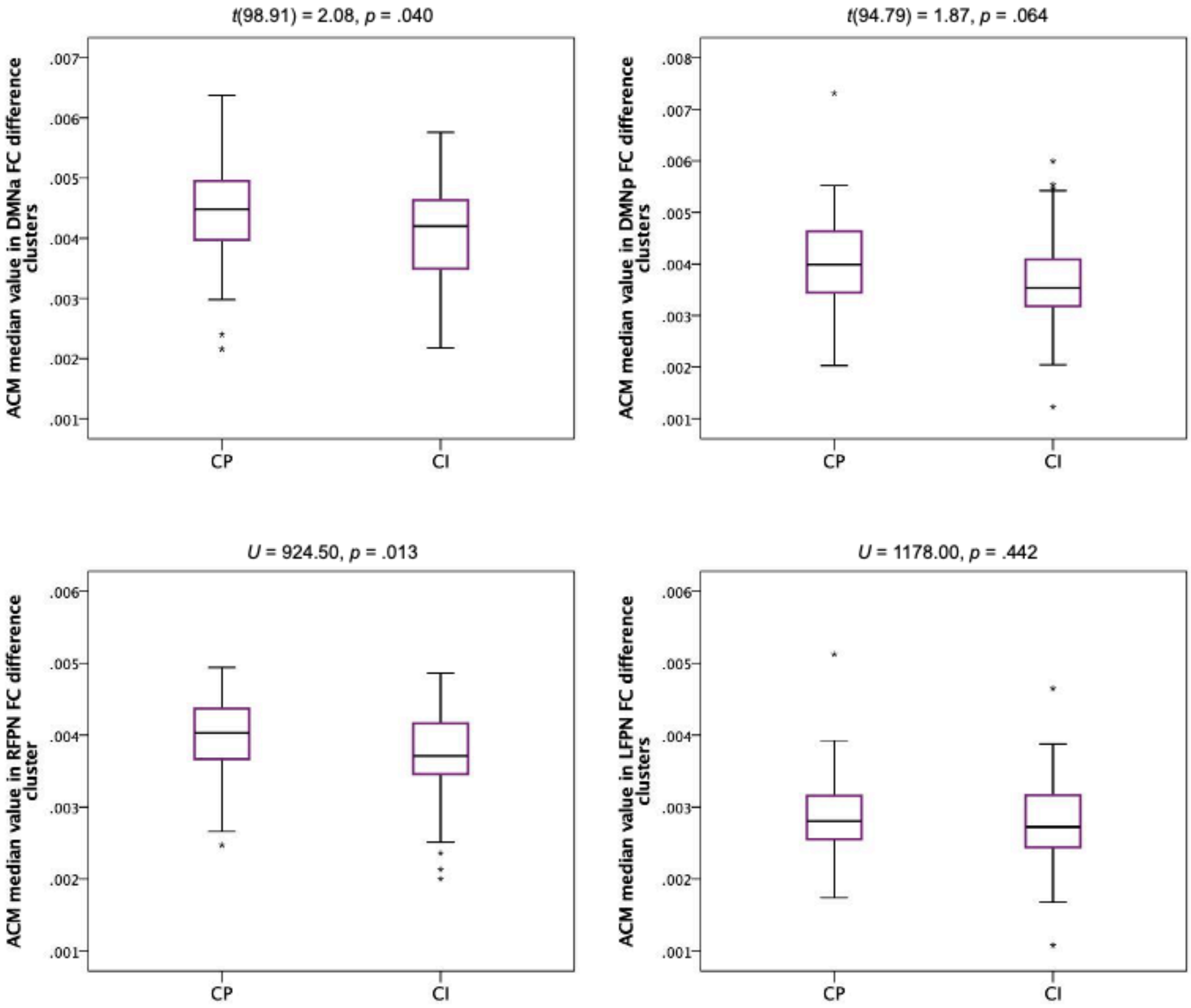

**B**

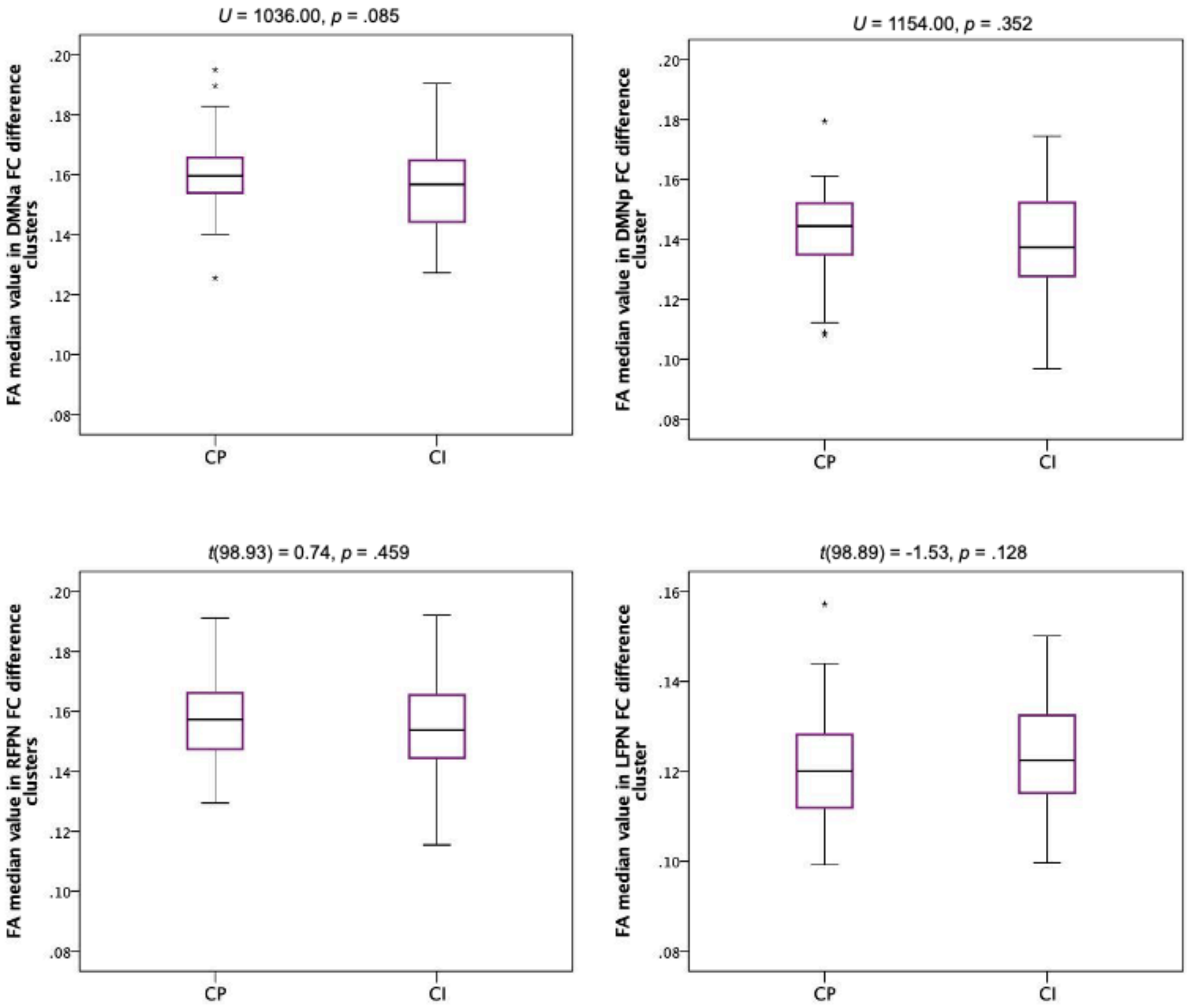
U = 1036.00, p = .085
FA median value in DMNa FC difference clusters
CP
CI
U = 1154.00, p = .352
FA median value in DMNp FC difference cluster
t(98.93) = 0.74, p = .459
FA median value in RFPN FC difference clusters
t(98.89) = -1.53, p = .128
FA median value in LFPN FC difference cluster

**C**

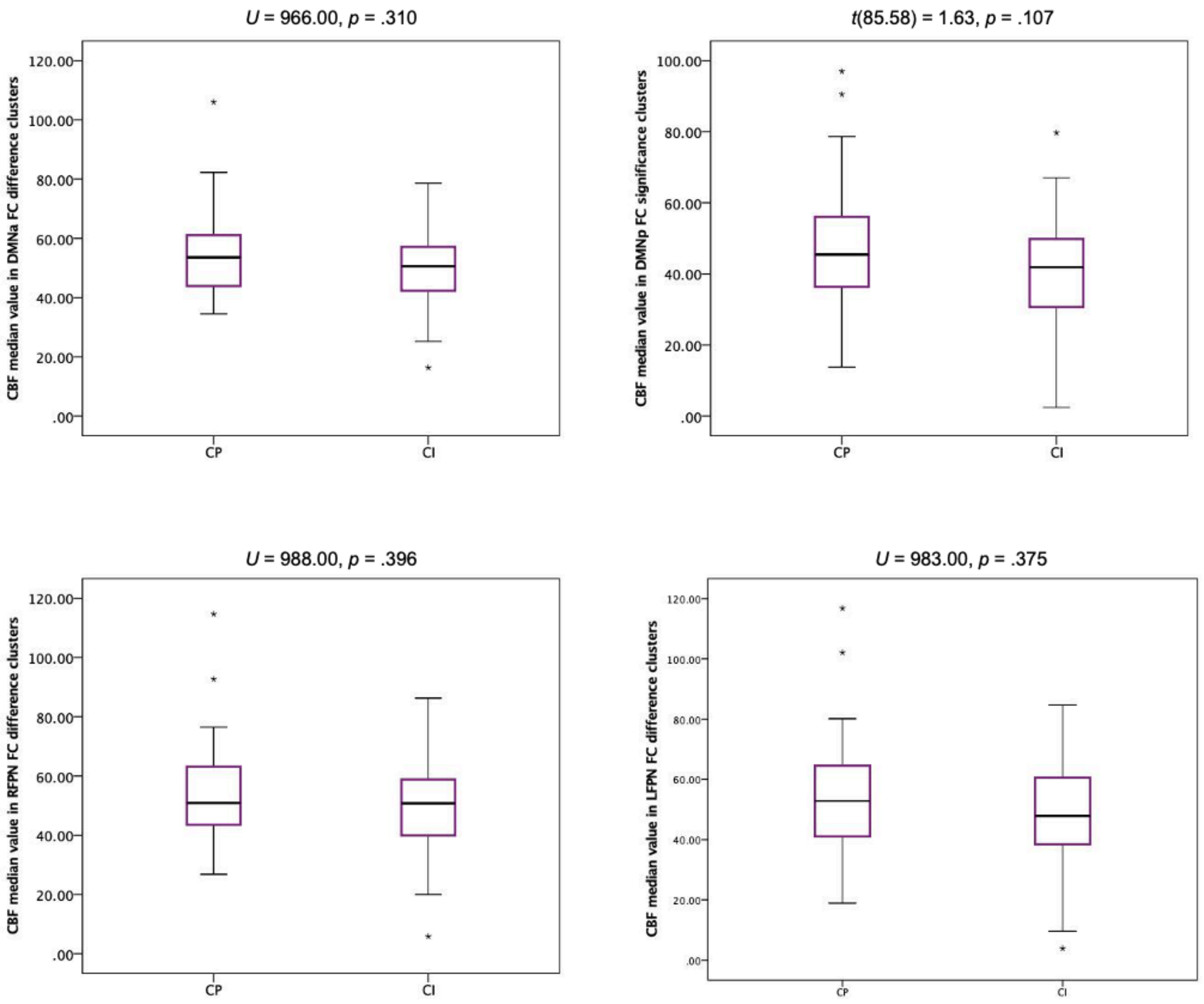


**Figure 2. Comparison of local ACM, FA and CBF changes in RSNs regions showing FC differences between CI and CP patients**

Figure 2 shows a comparison of median ACM values, normalised for head size (2a), median FA values (2b) and median CBF values (2c), between CI and CP patients, in the regions of the RSNs that showed FC changes in CI patients compared to CP patients. Boxes show median and interquartile range (IQL), and whiskers show range of values, excluding outliers. Stars indicate outliers. Results were considered significant at a Bonferroni corrected p threshold of $p \leq 0.0125$.

### Diffuse changes in connectivity and CBF within RSNs

Diffuse changes in connectivity and CBF were explored with two different approaches. We conducted region of interest analyses of ACM, FA and CBF spatial maps, as described. In addition, we masked these maps with the binarsed RSN masks and extracted the median ACM, FA and CBF values from each RSN. There are few papers in the multiple sclerosis literature on functional connectivity changes that have combined the rs-fMRI method with other MRI methods, and so there is a lack of consensus on the best approach for this. The voxelwise analysis approach can show the spatial location of any abnormalities in the metrics studied, but requires the abnormalities to be in the same spatial location within the ROI in most subjects in a group for a group difference to be detected. If this is not the case a group difference could be missed, so one alternative is to look at mean or median values of the metric within the ROI, which could detect differences is one group has abnormalities in the metric of interest compare to the other, even if these vary spatially across subjects.

We visually inspected the histograms of ACM, FA and CBF values in the RSN ROIs and as they were not normally distributed we extracted median, rather than mean, values.

Using this approach we found reduced ACM in the anterior DMN ($U$=897.00, $p$=0.008), in CI patients compared to CP patients, but no other RSNs. Similarly, there were no differences in median FA or CBF values in RSN regions between CI and CP patients (Fig. 3).

**A**

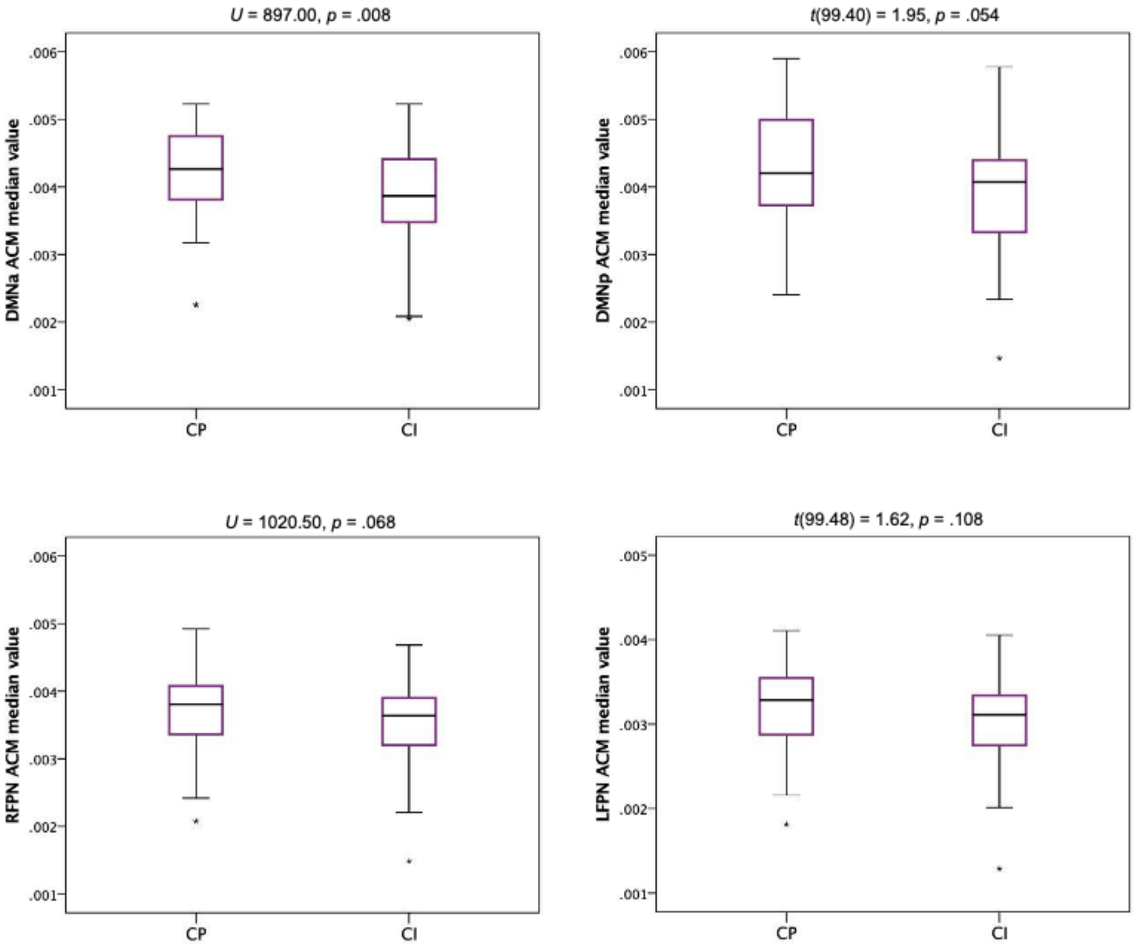

U = 897.00, p = .008
DMNa ACM median value
.006
.005
.004
.003
.002
.001
CP
CI
t(99.40) = 1.95, p = .054
DMNp ACM median value
.006
.005
.004
.003
.002
.001
CP
CI
U = 1020.50, p = .068
RFPN ACM median value
.006
.005
.004
.003
.002
.001
CP
CI
t(99.48) = 1.62, p = .108
LFPN ACM median value
.005
.004
.003
.002
.001
CP
CI

**B**

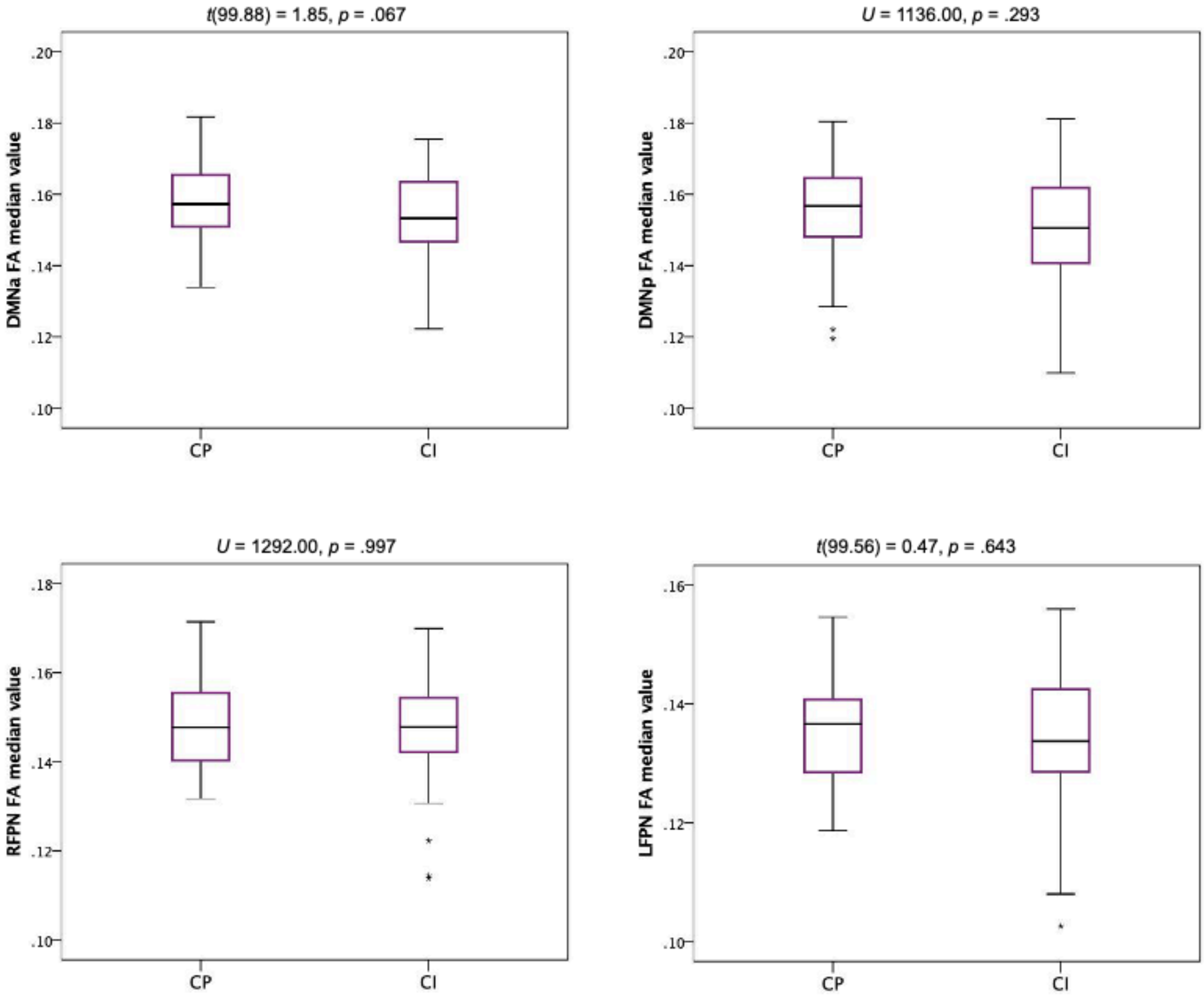

t(99.88) = 1.85, p = .067
DMNa FA median value
.20
.18
.16
.14
.12
.10
CP
CI
U = 1136.00, p = .293
DMNp FA median value
.20
.18
.16
.14
.12
.10
CP
CI
U = 1292.00, p = .997
RFPN FA median value
.18
.16
.14
.12
.10
CP
CI
t(99.56) = 0.47, p = .643
LFPN FA median value
.16
.14
.12
.10
CP
CI

**C**

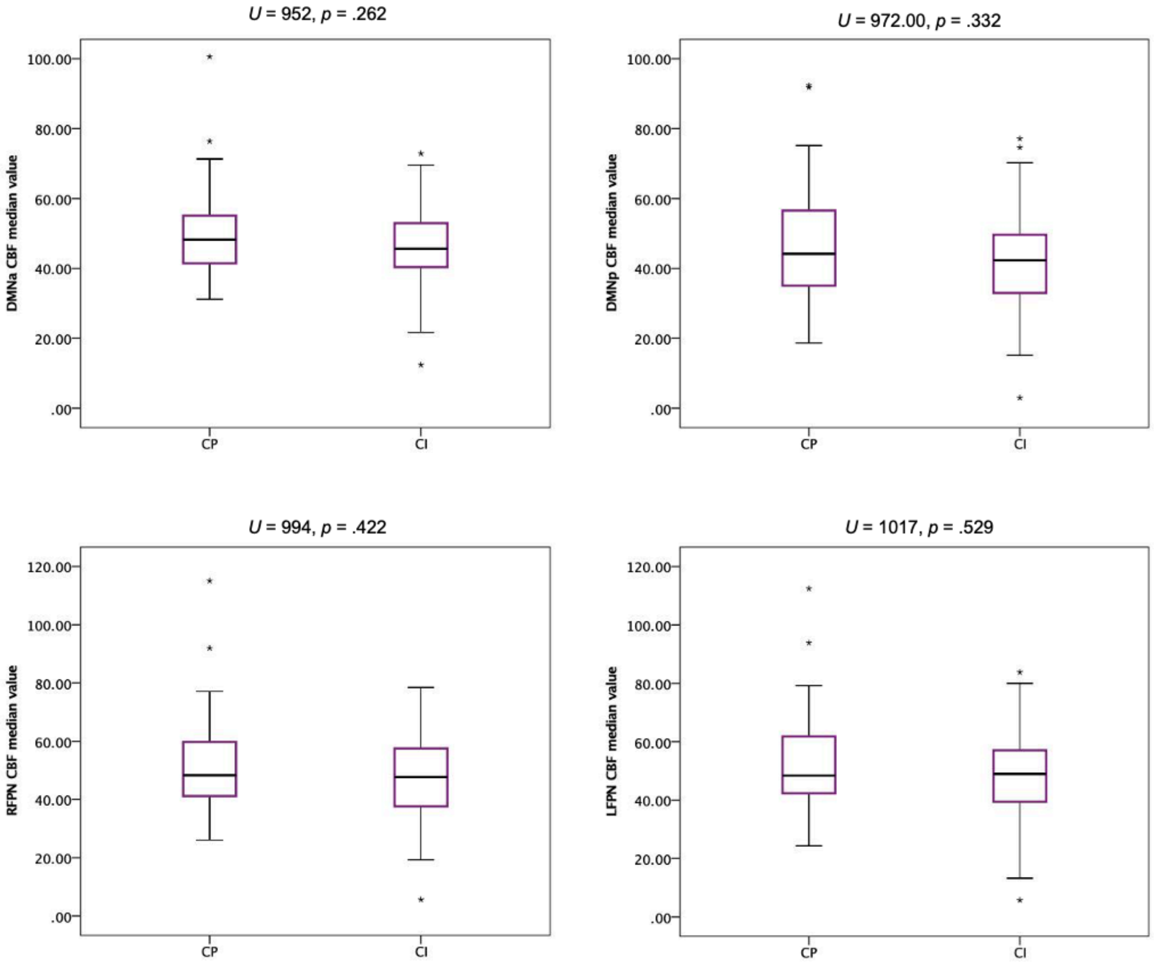


**Figure 3. Comparison of diffuse ACM, FA and CBF changes in RNSs between CI and CP patients**

Figure 3 shows a comparison of median ACM values, normalised for head size (3a), median FA values (3b) and median CBF values (3c), between CI and CP patients, throughout each RSN. Boxes show median and interquartile range (IQL), and whiskers show range of values, excluding outliers. Stars indicate outliers. Results were considered significant at a Bonferroni corrected p threshold of $p \leq 0.0125$

Voxelwise analyses of FA maps were conducted using two different approaches. First, we used tract-based spatial statistics (TBSS) (Smith *et al.*, 2006), as described. TBSS overcomes the difficulties of achieving accurate registration of the white matter (Soares *et al.*, 2013) by projecting all subjects' FA data onto a mean FA tract skeleton, before applying voxelwise cross-subject statistics. However, the FA skeleton created in TBSS includes only the centre of white matter tracts (Smith *et al.*, 2015) and may not capture white matter local to our functional network regions of interest, which are mainly in the grey matter. Therefore, we also conducted an exploratory voxelwise analysis of the non-skeletonised FA maps, by first registering these to MNI space using the same process as described for ACM maps and conducting permutation testing. Because of the limitations of each approach, and lack of similar prior studies in multiple sclerosis to direct our choice of method, we used both in an exploratory analysis, in order to gain an understanding of which is most suitable for an ROI analysis of RSNs.

Analyses of non-skeletonised FA maps were conducted in the same manner as the TBSS analysis, using an ROI approach to determine diffuse white matter changes within RSNs, and across the whole brain, to identify WM changes outside RSN regions.

The voxelwise analysis of FA maps found widespread FA reductions across the brain, as well as in each of the four RSNs of interest in CI patients (Fig. 4). FA changes were largely in the same locations as the FA changes identified from the TBSS analysis of skeletonised FA maps.

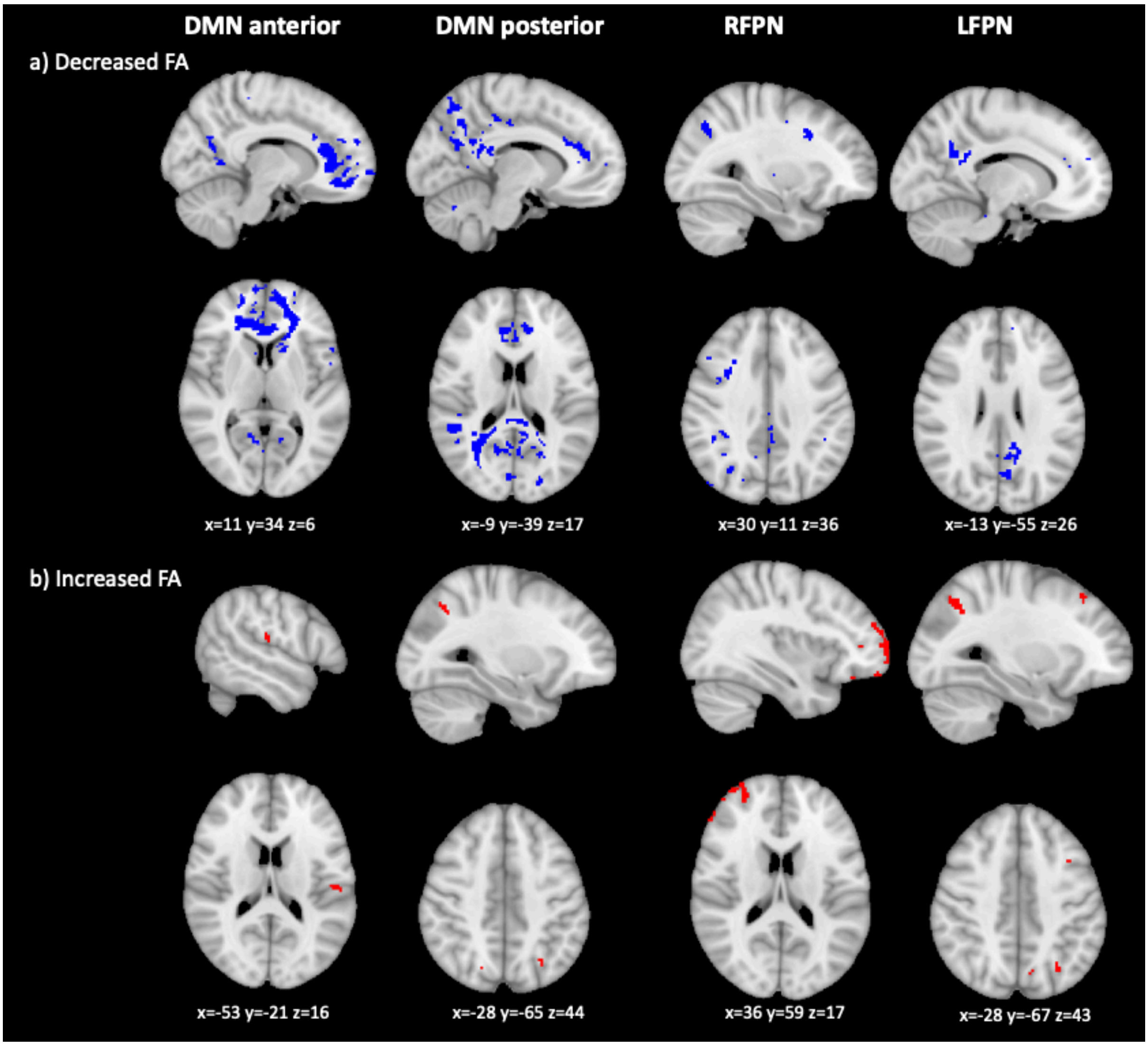

**Figure 4. FA changes in CI compared to CP patients, based on a voxelwise analysis of non-skeletonised FA maps**

Figure shows voxels showing FA abnormalities. Figure shows voxels showing ACM value abnormalities. Columns show each of the RSNs compared. The first row shows areas of decreased FA (in blue), the second row areas of increased FC (in red). MNI coordinates are given for the biggest voxel clusters displayed. Results were TFCE-corrected at $p \leq 0.05$, two-sided.

### Diffuse changes in connectivity and CBF throughout the brain – rationale and results

While our hypothesis is that we should see anatomical connectivity and CBF changes in RSNs that show FC changes in CI patients compared to CP patients, we also checked whether CI and CP groups showed differences in ACM, FA and CBF throughout the brain, by running voxelwise analyses without any ROI masks. This would help us understand our results better by determining the specificity of changes in the ACM, FA and CBF metrics to functional networks showing FC changes.

On ACM and FA from both TBSS and analysis of non-skeletonised maps we found widespread reductions throughout the brain in cognitively impaired compared to cognitively preserved patients (Fig. 5). On each analyses we found some voxels at the edges of the brain showing increases. Because of the small number of voxels and their location these are likely artifacts. Finally, the CBF analysis showed widespread CBF decreases, and no areas of CBF increases.

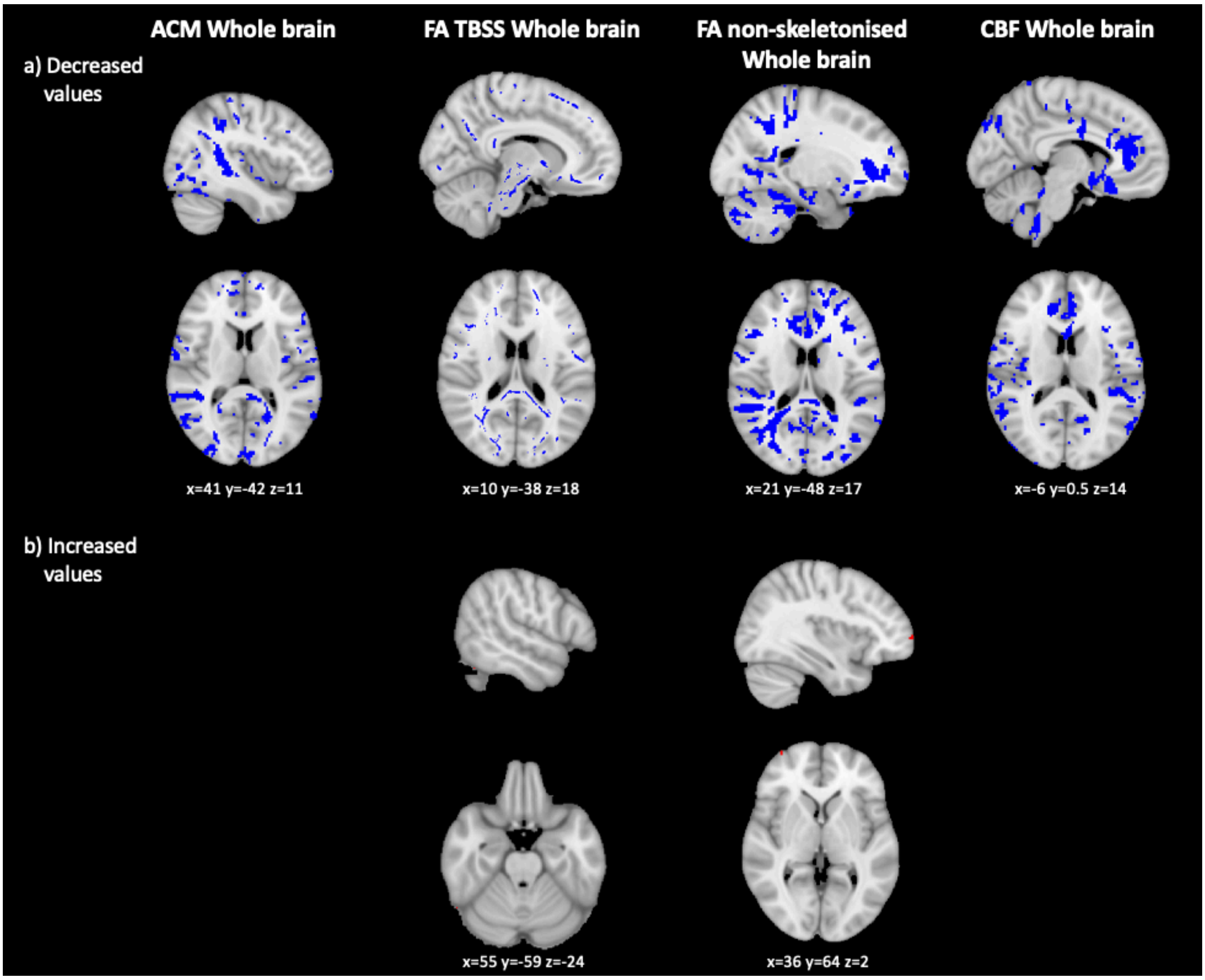


**Figure 5. Diffuse ACM, FA and CBF changes across the whole brain in CI compared to CP patients**

Figure shows ACM , FA and CBF abnormalities in. Columns show each of the metrics assessed: ACM, FA from TBSS, FA from analysis of non-skeletonised FA maps, and CBF, in that order. he first row shows areas of decreased values (in blue), the second row areas of increased values (in red). MNI coordinates are given for the biggest voxel clusters displayed. Results were TFCE-corrected at $p \leq 0.05$, two-sided
